# Hernia surgical treatment; multi-options and paucity of statistical conformation for the preferred surgical option

**DOI:** 10.1101/2022.12.28.22284003

**Authors:** Basheer Abdullah Marzoog, Kostin Sergey Vladimirovich

## Abstract

**Background:** Hernia is a common pathology in the globe and reported more frequently, particularly, inguinal hernia.

**Aims:** To identify the surgery of choice for the treatment of hernias by evaluating the required postoperative hospitalization time, as no other complications have been reported according to data from Mordovian Republic hospital.

**Material and methods:** A retrospective cohort study involved 790 patients for the period 2017-2022 treated surgically for various types of hernia; inguinal hernia, umbilical hernia, spontaneously reduced strangulated post-operative ventral hernia, incarcerated post-operative ventral hernia, and hernia of the Lina Alba. For statistical analysis, used T test, one way ANOVA test, and Pearson correlation test by using Statistica program.

**Results:** The hospitalization period after Lichtenstein surgery is statistically less than Postemsky surgery (mean 6.88 days, 7.43 days, respectively, t value -2.29593, p<0.02) and laparoscopic surgery (mean 6.88 days, 8.19072 days, respectively, t value 4,206817, p<0,000031). Whereas, Postemsky surgery has shorter post-operative hospitalization period in compare to laparoscopic surgery (mean 7.43 days, 8.19072 days, respectively, t value -2.19326, p<0.02). According to the used surgical approach, the patient’s post-operative hospitalization days differs (mean days: min. days; max. days, 7.50192: 0.00; 30.00). According to Postemsky (M ± m; 7.43262, ±0.167012), according to Martynov (M ± m; 8.37500, ±0.113440), according to Lichtenstein (M ± m; 6.88153, ±0.146845), according to Mayo (M ± m; 7.51282, ±0.280156), according to Bassini (M ± m; 8.77778, ± 2.379179), laparoscopically (M ± m; 8.19072, ± 0.268434), according to Sapezhko (M ± m; 8.25000, ± 1.380074), and another type of surgery (M ± m; 11.40000, ± 2.501999). Women (mean 8.525114 days) hospitalized longer than men (mean 7.065371 days), t value 5.871044, p< 0.001. A statistically significant correlation has been found between age and post-operative hospitalization time (Pearson Rank Order Correlations r=0.215561, p <0.05).

**Conclusions:** The study shows that the Lichtenstein surgery is the surgery of choice in terms of hospitalization time after the surgery. Straight association between sex and age with postoperative hospitalization days.

## Introduction

Hernia is a common pathology in the globe and remains paucity in the therapeutic techniques [1]. Management of such worldwide issue is crucial. Classically, hernias are classified as internal and external hernias. Each of which has its surgical approaches for management [1]. However, the statistical evidence on the surgery of choice requires more elaboration.

Hernia defined as a protrusion of the abdominal organs in another organ or through the abdominal wall as well as canals such as the inguinal canal [2]. In clinical practice, hernias are usually managed by strengthening the wall of the protruded tissue with mesh [3–5].

Postempsky approach involves complete elimination of the inguinal canal, the inguinal gap and in the creation of the inguinal canal with a completely new direction. The edge of the vagina of the rectus abdominis muscle, together with the connected tendon of the internal oblique and transverse muscles, is sewn to the upper pubic ligament. Next, the upper flap of aponeurosis, together with the internal oblique and transverse abdominal muscles, is sewn to the pubic-iliac cord and to the inguinal ligament. These sutures should push the spermatic cord to the lateral side to the limit. The lower flap of aponeurosis of the external oblique abdominal muscle, held under the spermatic cord, is fixed on top of the upper flap of aponeurosis. The newly formed “inguinal canal” with the spermatic cord should pass through the muscular-aponeurotic layer in an oblique direction from behind to front and from the inside to the outside so that its inner and outer openings are not opposite each other. The spermatic cord is placed on the aponeurosis and subcutaneous fat and skin are stitched over it [6].

Whereas, Martynov approach include sewn of the medial flap of the external oblique abdominal muscle to the inguinal ligament in front of the spermatic cord, and the lateral flap is sewn on top of the medial one. A duplicate is created from the flaps of dissected aponeurosis [6].

Lichtenstein method involves insertion of a polypropylene mesh of approximately 6×12 cm behind the spermatic cord. From below, it is sewn to the upper pubic and inguinal ligaments [7]. Laterally, it is cut, a “window” is made for the passage of the spermatic cord, then it is sewn again. It is sewn to the inner oblique and transverse muscles at the top, medially to the edge of the rectus muscle. Subsequently, the mesh sprouts connective tissue, and intraperitoneal pressure spreads evenly through it [6].

To date, there are more than 12 classifications, Nyhus classification, Gilbert classification, Aaachen classification [8, 9]. However, the European Hernia Society Board recommend using classification based on the Aachen system developed by Schumpelick et al. [10].

## Materials and methods

A retrospective cohort study involved 790 patients for the period 01.01.2017-28.02.2022 treated surgically for various types of hernia. The study involved 571 (72.28%) males and 219 (30.46%) females aged from 17-92 years old (mean; 59.23, standard error: 0.487868). 355 (44.93671%) patients live in the village and 435 (55.06329) live in the city.

Data collected from Mordovian Republic Hospital for the past 5 years and retrospectively analyzed. The consent of the patients has been taken for scientific purposes to analyze and publish the results of the study.

For statistical analysis, used T test, one way ANOVA test, and Pearson correlation test by using Statistica program (StatSoft, Inc. (2011). STATISTICA (data analysis software system), version 10. www.statsoft.com.).

## Results

The descriptive statistical analysis showed that the mean age of the male 58.06 years (minimum 17; maximum 92) years, whereas the mean age of the females 62.26941 (minimum 21; maximum 92) years. (Table 1) Approximately 8 different surgical approaches have been used to treat these patients and without any further life-threatening complications. According to Postemsky 141 (17.84810%) patient, according to Martynov 8 (1.01266 %) patients, according to Lichtenstein 287 (36.75%) patients, according to Mayo 117 (14.81013 %) patients, according to Bassini 9 (1.13924 %) patients, laparoscopically 194 (24.84 %) patients, patients, according to Sapezhko 20 (2.53165 %) patients, another way of plastic 5 (0.64%), and 9 (1.13924%) recovered on conservative therapy.

**Table 1:** Descriptive statistics of the sample.

| Variable | Descriptive Statistics |  |  |  |  |  |
| --- | --- | --- | --- | --- | --- | --- |
|  | Valid N | Mean | Minimum | Maximum | Std.Dev. | Standard Error |
| Total hospitalization days | 790 | 8,7608 | 1,0000 | 31,0000 | 3,21194 | 0,114276 |
| Age | 790 | 59,2278 | 17,0000 | 92,0000 | 13,71246 | 0,487868 |
| Ht1 | 790 | 41,8785 | 39,0000 | 50,0000 | 1,60020 | 0,056932 |
| Hb1 | 790 | 145,3380 | 115,0000 | 166,0000 | 10,94709 | 0,389480 |
| Le1 | 790 | 6,7265 | 3,1000 | 13,5000 | 1,90036 | 0,067612 |
| Er1 | 790 | 4,5978 | 3,7000 | 5,7000 | 0,50988 | 0,018141 |
| Er2 | 790 | 4,1978 | 3,3000 | 5,3000 | 0,50988 | 0,018141 |
| Hb2 | 790 | 144,3380 | 114,0000 | 165,0000 | 10,94709 | 0,389480 |
| Ht2 | 790 | 39,9759 | 37,0000 | 48,0000 | 1,64711 | 0,058602 |
| Le2 | 790 | 6,0765 | 2,6000 | 13,0000 | 1,61392 | 0,057421 |
| days before surgery | 785 | 1,3261 | 0,0000 | 7,0000 | 0,72713 | 0,025952 |
| Days after surgery | 785 | 7,4726 | 0,0000 | 30,0000 | 3,19032 | 0,113867 |

The mean age of patients who passed Martynov surgery is 58.75000, ST. Err. 5.793315. Mean age of patients who passed Lichtenstein surgery is 59.37282, ST. Err. 0.778549. Mean age of patients who have passed Mayo surgery is 59.08547, ST. Err. 1.287770. Mean age of patients who have passed Bassini surgery is 72.77778, ST. Err. 2.822419. Mean age of the patients who passed Laparoscopic surgery is 58.60309, ST. Err. 0.942875. Mean age of patients who passed Sapezhko surgery is 60.85000, ST. Err. 3.864327. Mean age of patients who passed Postemsky surgery is 58.46099, ST. Err. 1.242506. Mean age of patients who passed another type of surgery is 59.00000, ST. Err. 4.969909.

The hospitalization period after Lichtenstein surgery is statistically less than Postemsky surgery (mean 6.88 days, 7.43 days, respectively, t value -2.29593, p<0.02) and laparoscopic surgery (mean 6.88 days, 8.19072 days, respectively, t value 4,206817, p<0,000031). Whereas, laparoscopic surgery has longer post-operative hospitalization period in compare to Postemsky surgery (mean 8.19072 days, 7.43262 days, respectively, t value -2.19326, p<0.02). (Figure 1)

**Figure 1:**
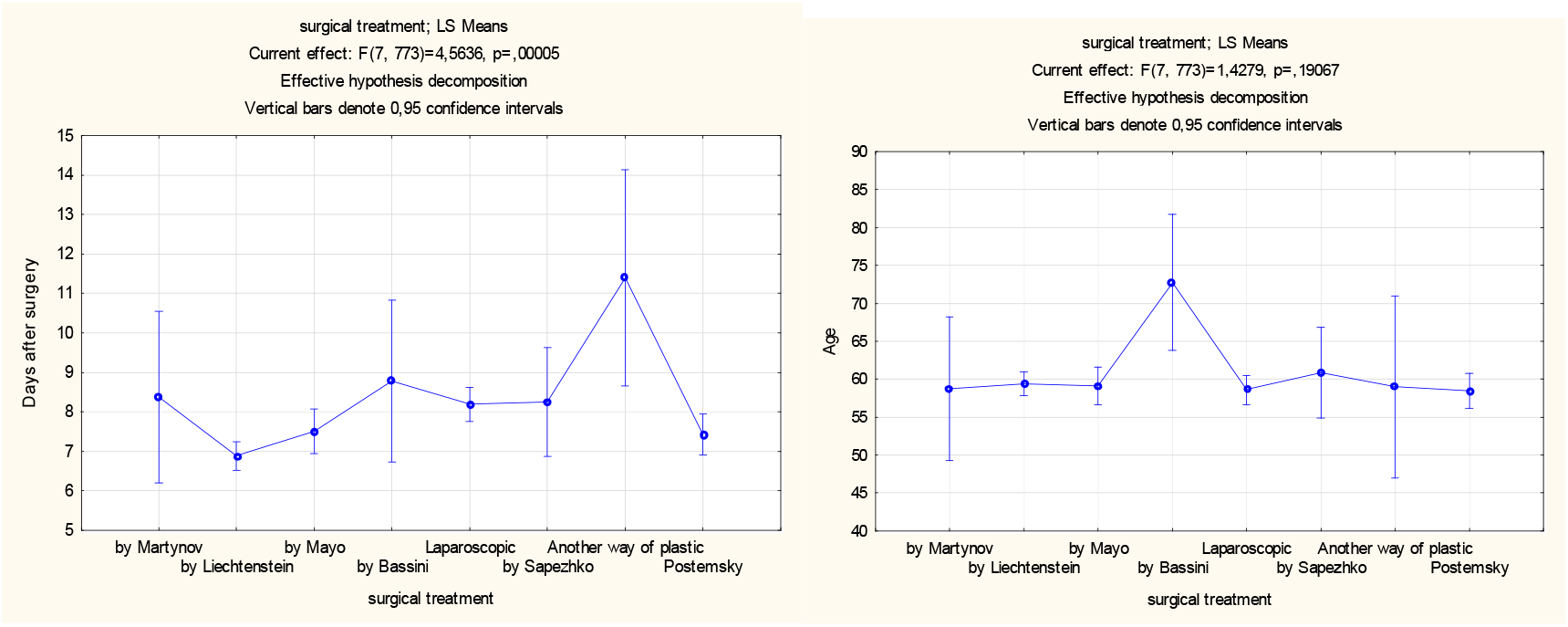
The shortest the postoperative hospitalization time is seen in Lichtenstein surgery and the longest in another way of plastic surgery (p< 0.00005). Age does not affect the choice of surgery (p=0.19067).

According to the used surgical approach, the patient’s post-operative hospitalization days differs (mean days: min. days; max. days, 7.50192: 0.00; 30.00). According to Postemsky (M ± m; 7.43262, ±0.167012), according to Martynov (M ± m; 8.37500, ±0.113440), according to Lichtenstein (M ± m; 6.88153, ±0.146845), according to Mayo (M ± m; 7.51282, ±0.280156), according to Bassini (M ± m; 8.77778, ± 2.379179), laparoscopically (M ± m; 8.19072, ± 0.268434), according to Sapezhko (M ± m; 8.25000, ± 1.380074), and another type of surgery (M ± m; 11.40000, ± 2.501999). (Table 2,3)

**Table 2:** The duration of post-operative hospitalization in different surgical approaches for hernia treatment.

|  |  |
| --- | --- |
|  | Descriptive Statistics |

| Effect | Level of Factor | N | Days after surgery Mean | Days after surgery Std.Dev. | Days after surgery St. Err | Days after surgery -95,00% | Days after surgery +95,00% |
| --- | --- | --- | --- | --- | --- | --- | --- |
| Total |  | 78<br>1 | 7,50192 | 3,170223 | 0,113440 | 7,279238 | 7,72460 |
| surgical treatment | by Martynov | 8 | 8,37500 | 5,180665 | 1,831642 | 4,043856 | 12,70614 |
| surgical treatment | by Lichtenstein | 28<br>7 | 6,88153 | 2,487713 | 0,146845 | 6,592499 | 7,17057 |
| surgical treatment | by Mayo | 11<br>7 | 7,51282 | 3,030350 | 0,280156 | 6,957936 | 8,06770 |
| surgical treatment | by Bassini | 9 | 8,77778 | 7,137538 | 2,379179 | 3,291381 | 14,26417 |
| surgical treatment | Laparoscopic | 19<br>4 | 8,19072 | 3,738848 | 0,268434 | 7,661282 | 8,72016 |
| surgical treatment | by Sapezhko | 20 | 8,25000 | 6,171880 | 1,380074 | 5,361471 | 11,13853 |
| surgical treatment | Another way of plastic | 5 | 11,40000 | 5,594640 | 2,501999 | 4,453337 | 18,34666 |
| surgical treatment | Postemsky | 14<br>1 | 7,43262 | 1,983161 | 0,167012 | 7,102432 | 7,76282 |

**Table 3:** Dependence of the postoperative hospitalization period on the type of surgery.

| Dependent:<br>Days after surgery | Median Test, Overall Median = 7,00000; Days after surgery<br>Independent (grouping) variable: surgical treatment<br>Chi-Square = 27,82642 df = 7 p = ,0002 |  |  |  |  |  |  |  |  |
| --- | --- | --- | --- | --- | --- | --- | --- | --- | --- |
|  | by Martynov | by Lichtenstein | by Mayo | by Bassini | Laparoscopic | by Sapezhko | Another way of plastic | Postemsky | Total |
| <= Median: observed | 6,00000 | 207,0000 | 66,0000 | 5,000000 | 101,0000 | 10,00000 | 1,00000 | 86,0000 | 482,0000 |
| expected | 4,93726 | 177,1242 | 72,2074 | 5,554417 | 119,7286 | 12,34315 | 3,08579 | 87,0192 |  |
| obs.-exp. | 1,06274 | 29,8758 | -6,2074 | -0,554417 | -18,7286 | -2,34315 | -2,08579 | -1,0192 |  |
| > Median: observed | 2,00000 | 80,0000 | 51,0000 | 4,000000 | 93,0000 | 10,00000 | 4,00000 | 55,0000 | 299,0000 |
| expected | 3,06274 | 109,8758 | 44,7926 | 3,445583 | 74,2714 | 7,65685 | 1,91421 | 53,9808 |  |
| obs.-exp. | -1,06274 | -29,8758 | 6,2074 | 0,554417 | 18,7286 | 2,34315 | 2,08579 | 1,0192 |  |
| Total: observed | 8,00000 | 287,0000 | 117,0000 | 9,000000 | 194,0000 | 20,00000 | 5,00000 | 141,0000 | 781,0000 |

The patients hospitalized before surgery for different periods ranged from zero days to 7 days (mean 1.3261, standard error 0.025952). Interestingly, gender plays important role in the determination of the post-operative period hospitalization time. Where women (mean 8.525114 days) hospitalized longer than men (mean 7.065371 days), t value 5.871044, p< 0.001.

Also, age plays important role in the hospitalization time, where statistically significant correlation has been found between age and post-operative hospitalization time (Spearman Rank Order Correlations r=0.215561, p <0.05). (Figure 2)

**Figure 2:**
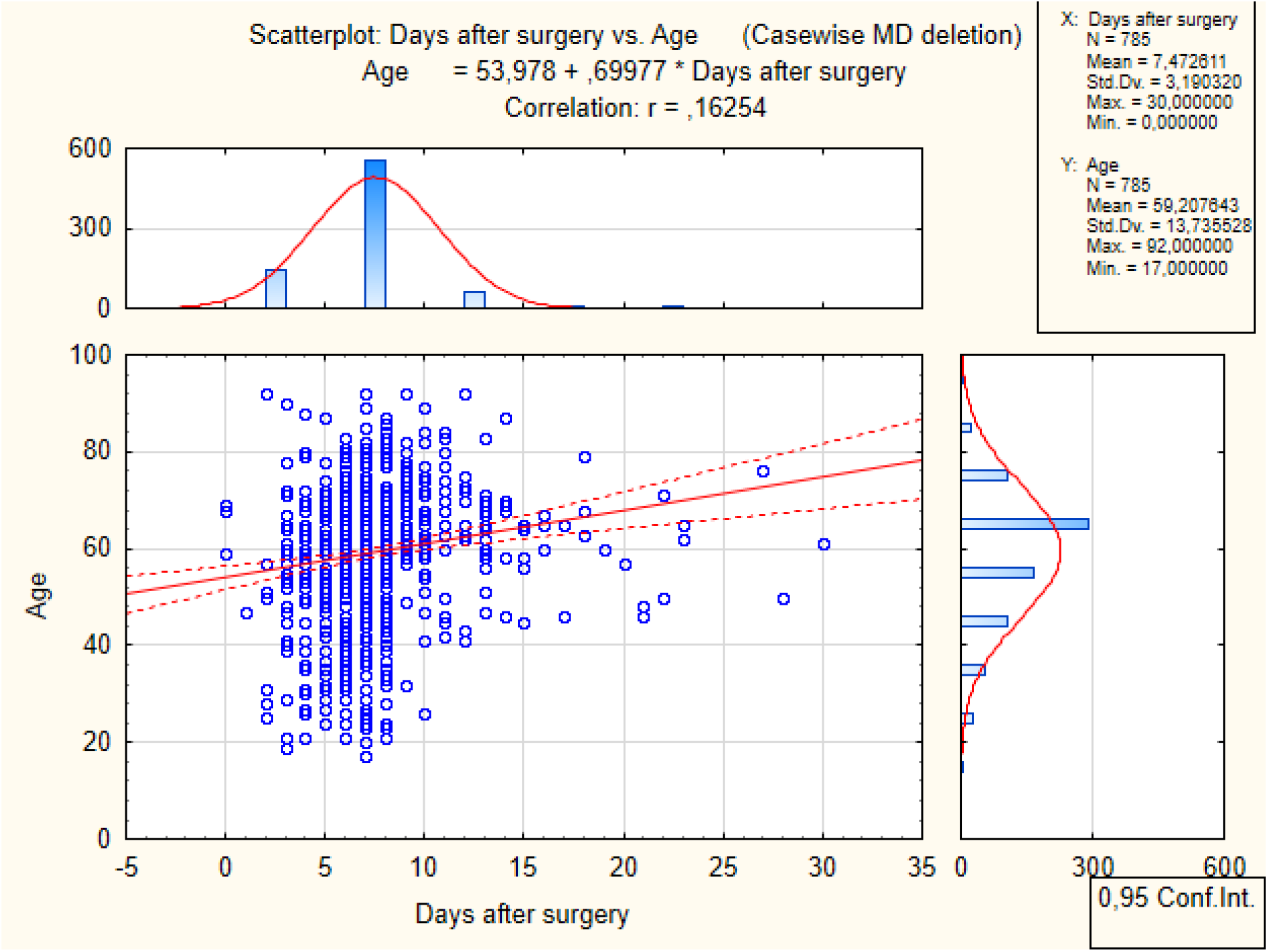
Elderly people have longer post-operative hospitalization time.

Since Severe acute respiratory syndrome-Corona Virus appeared in the late 2019, not all patients have been checked for the association between corona virus infection disease -19 (COVID-19) and the post-operative hospitalization period. Out of 485 patients, only 51 (8.72%) are having antibodies against COVID-19 infection. There is no statistically significant correlation between the COVID-19 infection and the post-operative hospitalization period.

A statistically significant association has been found between erythrocyte level and the post-operative hospitalization period. Data did not show a relationship between the home and the postoperative hospitalization period.

By the localization and type of the hernia, the most frequently reported localization is post-operative ventral hernia 86 (10.96%) patients of the total sample. These patients have an average hospitalization time 10.05814 days, St. err. ± 0.409983) and an average age of 61.84884, St. err. 1.14054. Approximately 72 different types of hernia have been reported. (Table 4)

**Table 4:** The incidence rate of different types of hernias and there required post-operative hospitalization days in Mordovia Republic. Abbreviations: p/o; post-operative.

| Effect | Descriptive Statistics |  |  |  |  |  |  |
| --- | --- | --- | --- | --- | --- | --- | --- |
|  | Level of Factor | N | Days after surgery Mean | Days after surgery Std.Dev. | Days after surgery St. Err | Days after surgery -95,00% | Days after surgery +95,00% |
| Total |  | 785 | 7,47261 | 3,19032 | 0,113867 | 7,2491 | 7,69613 |
| Hernia localization | 1. Right-sided direct inguinal hernia | 56 | 6,85714 | 1,63405 | 0,218360 | 6,4195 | 7,29475 |
| Hernia localization | 2. Strangulated right inguinal hernia | 14 | 8,21429 | 2,15473 | 0,575876 | 6,9702 | 9,45839 |
| Hernia localization | 3. Left side oblique inguinal hernia | 27 | 6,51852 | 1,50308 | 0,289269 | 5,9239 | 7,11312 |
| Hernia localization | 4. Right-sided oblique inguinal hernia | 30 | 6,90000 | 1,15520 | 0,210909 | 6,4686 | 7,33136 |
| Hernia localization | 5. Right-sided oblique inguinal hernia | 71 | 6,21127 | 1,73135 | 0,205474 | 5,8015 | 6,62107 |
| Hernia localization | 6. Hernia of the white line of the abdomen | 23 | 7,65217 | 2,69020 | 0,560945 | 6,4888 | 8,81550 |
| Hernia localization | 7. P/o ventral hernia | 86 | 10,05814 | 3,80203 | 0,409983 | 9,2430 | 10,87330 |
| Hernia localization | 8. Giant p/o ventral hernia | 1 | 5,00000 |  |  |  |  |
| Hernia localization | 9. Strangulated hernia of the white line of the abdomen | 9 | 6,11111 | 2,14735 | 0,715783 | 4,4605 | 7,76171 |
| Hernia localization | 10. Strangulated umbilical hernia | 28 | 6,89286 | 2,57249 | 0,486156 | 5,8953 | 7,89037 |
| Hernia localization | 11. Right femoral hernia | 2 | 8,00000 | 1,41421 | 1,000000 | -4,7062 | 20,70620 |
| Hernia localization | 12. P / o ventral hernia of large sizes | 2 | 14,50000 | 6,36396 | 4,500000 | -42,6779 | 71,67792 |
| Hernia localization | 13. Right-sided direct inguinal hernia | 46 | 6,89130 | 2,66857 | 0,393459 | 6,0988 | 7,68377 |
| Hernia localization | 14. Umbilical hernia | 61 | 6,65574 | 2,56830 | 0,328837 | 5,9980 | 7,31351 |
| Hernia localization | 15. Incarcerated p/o ventral hernia | 33 | 9,06061 | 4,09984 | 0,713691 | 7,6069 | 10,51435 |
| Hernia localization | 16. Spontaneously reduced strangulated umbilical hernia | 7 | 5,14286 | 2,11570 | 0,799660 | 3,1862 | 7,09955 |
| Hernia localization | 17. Spontaneously reduced p/o ventral hernia | 3 | 9,66667 | 4,61880 | 2,666667 | -1,8071 | 21,14041 |
| Hernia localization | 18. Left-sided oblique inguinal hernia | 61 | 6,75410 | 3,34493 | 0,428274 | 5,8974 | 7,61077 |
| Hernia localization | 19. Strangulated right femoral hernia | 3 | 6,00000 | 3,00000 | 1,732051 | -1,4524 | 13,45241 |
| Hernia localization | 20. Strangulated left femoral hernia | 3 | 14,00000 | 11,35782 | 6,557439 | -14,2144 | 42,21438 |
| Hernia localization | 21. Strangulated spontaneously reduced umbilical hernia | 2 | 7,00000 | 2,82843 | 2,000000 | -18,4124 | 32,41241 |
| Hernia localization | 22. Incarcerated spontaneously reduced left-sided inguinal hernia | 3 | 7,00000 | 1,00000 | 0,577350 | 4,5159 | 9,48414 |
| Hernia localization | 23. Axial hiatal hernia | 5 | 10,40000 | 6,46529 | 2,891366 | 2,3723 | 18,42772 |
| Hernia localization | 24. Recurrent p/o ventral hernia | 11 | 9,90909 | 2,58668 | 0,779913 | 8,1713 | 11,64685 |
| Hernia localization | 25. Spontaneously reduced strangulated hernia of the white line of the abdomen | 2 | 5,00000 | 1,41421 | 1,000000 | -7,7062 | 17,70620 |
| Hernia localization | 26. Spontaneously right restrained p / o ventral hernia | 4 | 9,75000 | 2,50000 | 1,250000 | 5,7719 | 13,72806 |
| Hernia localization | 27. Recurrent umbilical hernia | 2 | 8,50000 | 6,36396 | 4,500000 | -48,6779 | 65,67792 |
| Hernia localization | 28. Recurrent hernia of the linea alba | 2 | 8,50000 | 3,53553 | 2,500000 | -23,2655 | 40,26551 |
| Hernia localization | 29. Spontaneously reduced strangulated p/o ventral hernia | 3 | 6,66667 | 5,13160 | 2,962731 | -6,0809 | 19,41427 |
| Hernia localization | 30. Recurrent strangulated p/o ventral hernia | 1 | 30,00000 |  |  |  |  |
| Hernia localization | 31. Strangulated right-sided direct inguinal hernia | 16 | 7,75000 | 4,71169 | 1,177922 | 5,2393 | 10,26068 |
| Hernia localization | 32. Left femoral hernia | 1 | 4,00000 |  |  |  |  |
|  | Level of Factor | N | Days after surgery Mean | Days after surgery Std.Dev. | Days after surgery St. Err | Days after surgery -95,00% | Days after surgery +95,00% |
| Hernia localization | 33. Spontaneously reduced strangulated umbilical hernia | 1 | 6,00000 |  |  |  |  |
| Hernia localization | 34. Left direct inguinal hernia | 22 | 7,36364 | 1,36436 | 0,290882 | 6,7587 | 7,96856 |
| Hernia localization | 35. Strangulated right-sided inguinal-scrotal hernia | 3 | 7,00000 | 1,00000 | 0,577350 | 4,5159 | 9,48414 |
| Hernia localization | 36. Recurrent left-sided inguinal hernia | 1 | 8,00000 |  |  |  |  |
| Hernia localization | 37. Strangulated recurrent right inguinal hernia | 2 | 7,50000 | 0,70711 | 0,500000 | 1,1469 | 13,85310 |
| Hernia localization | 38. Recurrent left-sided inguinal hernia | 15 | 7,00000 | 1,64751 | 0,425385 | 6,0876 | 7,91236 |
| Hernia localization | 39. Strangulated left inguinal hernia | 6 | 7,33333 | 0,51640 | 0,210819 | 6,7914 | 7,87526 |
| Hernia localization | 40. Right-sided inguinal-scrotal hernia | 18 | 8,61111 | 3,68046 | 0,867492 | 6,7809 | 10,44136 |
| Hernia localization | 41. Left-sided inguinal-scrotal hernia | 5 | 7,40000 | 2,19089 | 0,979796 | 4,6797 | 10,12035 |
| Hernia localization | 42. Left side oblique inguinal hernia | 4 | 7,25000 | 0,50000 | 0,250000 | 6,4544 | 8,04561 |
| Hernia localization | 43. Left-sided oblique inguinal-scrotal hernia | 3 | 6,66667 | 0,57735 | 0,333333 | 5,2324 | 8,10088 |
| Hernia localization | 44. Left-sided sliding inguinal hernia | 5 | 6,60000 | 1,14018 | 0,509902 | 5,1843 | 8,01571 |
| Hernia localization | 45. Left-sided sliding inguinal-scrotal hernia | 1 | 7,00000 |  |  |  |  |
| Hernia localization | 46. Recurrent right inguinal hernia | 7 | 6,71429 | 1,60357 | 0,606092 | 5,2312 | 8,19734 |
| Hernia localization | 47. Right sided inguinal hernia | 1 | 9,00000 |  |  |  |  |
| Hernia localization | 48. Recurrent right-sided inguinal-scrotal hernia | 3 | 7,66667 | 1,52753 | 0,881917 | 3,8721 | 11,46125 |
| Hernia localization | 49. Spontaneously reduced strangulated right inguinal hernia | 1 | 4,00000 |  |  |  |  |
| Hernia localization | 50. Spontaneously reduced strangulated left inguinal hernia | 1 | 8,00000 |  |  |  |  |
| Hernia localization | 51. Recurrent right inguinal hernia | 1 | 5,00000 |  |  |  |  |
| Hernia localization | 52. Strangulated left inguinal hernia | 15 | 5,86667 | 2,16685 | 0,559478 | 4,6667 | 7,06663 |
| Hernia localization | 53. Recurrent right-sided direct inguinal hernia | 5 | 6,80000 | 0,83666 | 0,374166 | 5,7611 | 7,83885 |
| Hernia localization | 54. Left-sided direct inguinal hernia | 17 | 6,41176 | 2,52633 | 0,612725 | 5,1128 | 7,71068 |
| Hernia localization | 55. Recurrent left-sided strangulated inguinal hernia | 1 | 7,00000 |  |  |  |  |
| Hernia localization | 56. Right-sided sliding inguinal hernia | 2 | 8,50000 | 0,70711 | 0,500000 | 2,1469 | 14,85310 |
| Hernia localization | 57. Strangulated recurrent inguinal-scrotal hernia | 1 | 8,00000 |  |  |  |  |
| Hernia localization | 58. Incarcerated spontaneously reduced right-sided direct inguinal hernia line | 2 | 5,50000 | 0,70711 | 0,500000 | -0,8531 | 11,85310 |
| Hernia localization | 59. Strangulated recurrent Right-sided direct inguinal hernia line | 1 | 6,00000 |  |  |  |  |
| Hernia localization | 60. Sliding left inguinal hernia | 1 | 6,00000 |  |  |  |  |
| Hernia localization | 61. Strangulated sliding left inguinal hernia | 1 | 6,00000 |  |  |  |  |
| Hernia localization | 62. Spontaneously reduced strangulated left-sided inguinal hernia | 1 | 8,00000 |  |  |  |  |
| Hernia localization | 63. Spontaneously reduced strangulated left inguinal hernia | 3 | 8,00000 | 3,46410 | 2,000000 | -0,6053 | 16,60531 |
| Hernia localization | 64. Incarcerated left-sided inguinal-scrotal hernia | 5 | 9,00000 | 6,55744 | 2,932576 | 0,8579 | 17,14214 |
| Hernia localization | 65. Left-sided recurrent sliding inguinal hernia | 1 | 10,00000 |  |  |  |  |
| Hernia localization | 66. Right-sided oblique inguinal-scrotal hernia | 1 | 7,00000 |  |  |  |  |
| Hernia localization | 67. Spontaneously reduced strangulated right-sided sliding inguinal-scrotal hernia | 1 | 7,00000 |  |  |  |  |
| Hernia localization | 68. Spontaneously reduced strangulated Right-sided direct inguinal hernia line | 4 | 5,50000 | 1,91485 | 0,957427 | 2,4530 | 8,54696 |
|  | Level of Factor | N | Days after surgery Mean | Days after surgery Std.Dev. | Days after surgery St. Err | Days after surgery -95,00% | Days after surgery +95,00% |
| Hernia localization | 69. Bilateral inguinal hernia | 1 | 6,00000 |  |  |  |  |
| Hernia localization | 70. Recurrent strangulated right-sided direct inguinal hernia line | 2 | 5,00000 | 1,41421 | 1,000000 | -7,7062 | 17,70620 |
| Hernia localization | 71. Recurrent bilateral inguinal hernia | 1 | 9,00000 |  |  |  |  |
| Hernia localization | 72. Spontaneously reduced strangulated right-sided inguinal-scrotal hernia | 1 | 10,00000 |  |  |  |  |

Patients have statistically significant longer postoperative hospitalization days in case of post-operative ventral hernia and strangulated umbilical hernia (t value 4.103840, p=0.000077). Also, have statistically significant longer post-operative hospitalization days in case of post-operative ventral hernia and umbilical hernia (t value 6.072506, p=0.000000).

Depending on the age, the risk of different hernias increases. The most common hernia for young people is recurrent right-sided inguinal-scrotal hernia (mean age by years 40.66667, St. Er. 7.68838). For elderly people, most common type of hernia is strangulated left femoral hernia (mean age by years 78.66667, St. Er. 7.05534). The mean age for hernia development in both men and female is 59.22785 years, St. Err. 0.48787.

Choosing the surgery type is a hernia type dependent and age dependent. The mean age of the patient who passed Martynov surgery was 58.75000, St. Er. 5.793315. Mean age of patient passed Lichtenstein surgery 59.37282, St. Er. 0.778549. The mean age of the patient passed Mayo surgery 59.08547, St. Er. 1.287770. Mean age of patient passed Bassini surgery 72.77778, St. Er. 2.822419. Mean age for patient passed Laparoscopic surgery 58.60309, St. Er. 0.942875. Mean age of patient passed Sapezhko surgery 60.85000, St. Er. 3.864327. The mean age of the patient passed Postemsky surgery 58.46099, St. Er. 1.242506. Mean age of patient passed another type of plastic surgery 59.00000, St. Er. 4.969909.

### Current prospective for future therapeutic strategy

The current advancement in the surgical treatment has been achieved huge advances in improving the prognosis, reduce hospitalization days and less complications frequency associated with surgery [5]. The current approaches for treatment of the most frequently reported hernia remains poorly developed and requires further investigations [11]. Using of transabdominal preperitoneal (TAPP) repair and totally extraperitoneal (TEP) surgery for treatment of inguinal hernia is safe and effective method [12]. According to a systemic review results, the TEP has longer surgery time, shorter total hospitalization days, earlier discharge [13]. The recurrence rates of hernia in post TEP are similar to those for open inguinal hernia repair [13]. However, the TEP involves greater expenses for hospitals, but appears to be cost effective from a societal perspective [13].

Recent study demonstrated that applying combined mesh repair with autologous tissue repair has more efficacy in preventing the recurrence of inguinal hernia [14]. The current guidelines for hiatal hernias strongly indicated repair of symptomatic paraoesophageal hiatal hernias, particularly those with acute obstructive symptoms or which have undergone volvulus [15].

## Discussion

In our country, the standards of medical care until December 2019 included the standard one day of hospitalization before surgery for laboratory tests and the postoperative period until the removal of the skin sutures or stable dynamics of the wound process. Therefore, patients were not discharged for outpatient treatment with the presence of skin sutures. The criterion of complete recovery was considered to be complete healing of the skin at the site of surgery. The time for removing stitches from the skin is 7-10 days. Whereas, in plastic surgery this criterion does not exist. With hernioplasty, local tissues are sewn together and the faster they fuse, the better the result. During laparoscopy, the local tissues do not participate in the operation. Thus, the difference in the terms of hospitalization characterizes only the methods of surgical intervention (open and laparoscopic; plastic surgery with local tissues and mesh graft, strengthening of the anterior and posterior walls of the inguinal canal).

Males at an early age are affected in hernia more than females. Also, the incidence rate of hernia is seen more frequently in male than in female. The choice of surgical treatment method depends on the localization of the hernia, the risk of complications development, and the postoperative recovery time as well as the choice of the patient [4, 7, 22, 11, 14, 16–21]. Also, with age hernia risk increases. Our statistical data constant with the previous recommendation of the American College of Surgeons. Where they recommend using Lichtenstein hernia repair [7]. Several advantages are in favor of Lichtenstein surgery including the low hernia recurrence rate, low risk of complications, ability to perform in outpatient manner [20, 23]. We add to that Lichtenstein surgery has the less recovery days after surgery.

The recurrence of hernia has been assessed by a randomized clinical trial and showed no statistical difference between laparoscopic and Lichtenstein surgery [24]. However, some other meta-analysis showed superiority of laparoscopic procedure on Lichtenstein in terms of patient satisfaction [25].

Inguinal hernias remain the most commonly reported hernia worldwide. Annually, 20 million patients treat surgically inguinal hernia. Post-operative sequelae are also crucial in terms social life and daily activity. Using the Short Form-36 is (SF-36) is an acceptable tool to assesses the post-operative patient’s health status. Additionally, development of severe pain in some patients has been reported in some patients, which is also reported in other studies [26].

## Conclusions

In the light of our results, statistically significant correlation between the type of the surgery and the post-operative period hospitalization days have been identified. Also, our study showed that Lichtenstein surgery is the surgery of choice in terms of the hospitalization time after the surgery. Straight association between sex and age with postoperative hospitalization days. The relationship between laboratory values and postoperative hospitalization time shows a poor correlation.

## Data Availability

All data produced in the present study are available upon reasonable request to the authors

## Declarations

Authors’ contributions: MB analyzed the statistical data, wrote the draft, and revised the final version of the paper, KS collected the data from the hospital. All authors have read and approved the manuscript.

### List of abbreviations

COVID-19: corona virus infection disease -19,
TAPP: transabdominal preperitoneal,
TEP: totally extraperitoneal

## ETHICS APPROVAL AND CONSENT TO PARTICIPATE

The study approved by the National Research Mordovia State University, Russia, from “Ethics Committee Requirement N8/2 from 30.06.2021”.

## HUMAN AND ANIMAL RIGHTS

No animals were used in this research. All human research procedures followed were in accordance with the ethical standards of the committee responsible for human experimentation (institutional and national), and with the Helsinki Declaration of 1975, as revised in 2013.

## CONSENT FOR PUBLICATION

Written informed consent was obtained from the patients for publication of study results and any accompanying images.

## STANDARDS OF REPORTING

STROBE guideline has been followed.

## AVAILABILITY OF DATA AND MATERIALS

Not applicable.

## ACKNOWLEDGEMENTS

Declared none.

## References

[1] International Guidelines for Groin Hernia Management. Hernia, 2018, 22 (1), 1–165. https://doi.org/10.1007/s10029-017-1668-x.

[2] Grove, T. N.; Kontovounisios, C.; Montgomery, A.; Heniford, B. T.; Windsor, A. C. J.; Warren, O. J.; de Beaux, A.; Boermeester, M.; Bougard, H.; Butler, C.; et al. Perioperative Optimization in Complex Abdominal Wall Hernias: Delphi Consensus Statement. BJS Open, 2021, 5 (5). https://doi.org/10.1093/bjsopen/zrab082.

[3] Aguirre, D. A.; Santosa, A. C.; Casola, G.; Sirlin, C. B. Abdominal Wall Hernias: Imaging Features, Complications, and Diagnostic Pitfalls at Multi-Detector Row CT. Radiographics, 2005, 25 (6), 1501–1520. https://doi.org/10.1148/RG.256055018/ASSET/IMAGES/LARGE/G05NV01G30B.JPEG.

[4] Vitous, C. A.; Jafri, S. M.; Seven, C.; Ehlers, A. P.; Englesbe, M. J.; Dimick, J.; Telem, D. Exploration of Surgeon Motivations in Management of Abdominal Wall Hernias: A Qualitative Study. JAMA Netw. open, 2020, 3 (9), e2015916. https://doi.org/10.1001/jamanetworkopen.2020.15916.

[5] Andresen, K.; Rosenberg, J. [Development in Abdominal Hernia Repair]. Ugeskr. Laeger, 2016, 178 (30).

[6] Кузин, М. И.; Шкроб, О. С.; Кузин, Н. М.; КРЫЛОВ, Н. Н.; УСПЕНСКИЙ, Л. В.; КУЛАКОВА, А. М.; АРТЮХИНА, Е. Г.; ЧИСТОВ, Л. В.; ШКРОБ, О. С. Хирургические Болезни, 3rd ed.; Кузина, М. И., Ed.; Медицина, 2002.

[7] Amid, P. K. Lichtenstein Tension-Free Hernioplasty: Its Inception, Evolution, and Principles. Hernia, 2004, 8 (1), 1–7. https://doi.org/10.1007/s10029-003-0160-y.

[8] Gilbert, A. I. An Anatomic and Functional Classification for the Diagnosis and Treatment of Inguinal Hernia. Am. J. Surg., 1989, 157 (3), 331–333. https://doi.org/10.1016/0002-9610(89)90564-3.

[9] Schumpelick, V.; Treutner, K. H.; Arlt, G. [Classification of Inguinal Hernias]. Chirurg., 1994, 65 (10), 877–879.

[10] Kulacoglu, H.; Ozdogan, M.; Gurer, A.; Ersoy, E. P.; Onder Devay, A.; Duygulu Devay, S.; Gulbahar, O.; Gogkus, S. Prospective Comparison of Local, Spinal, and General Types of Anaesthesia Regarding Oxidative Stress Following Lichtenstein Hernia Repair. Bratisl. Lek. Listy, 2007, 108 (8), 335–339. https://doi.org/18203536.

[11] Knaapen, L.; Buyne, O.; Slater, N.; Matthews, B.; Goor, H.; Rosman, C. Management of Complex Ventral Hernias: Results of an International Survey. BJS Open, 2021, 5 (1). https://doi.org/10.1093/bjsopen/zraa057.

[12] Cao, C.; Shi, X.; Jin, W.; Luan, F. Clinical Data Analysis for Treatment of Adult Inguinal Hernia by TAPP or TEP. Front. Surg., 2022, 9. https://doi.org/10.3389/fsurg.2022.900843.

[13] Kuhry, E.; van Veen, R. N.; Langeveld, H. R.; Steyerberg, E. W.; Jeekel, J.; Bonjer, H. J. Open or Endoscopic Total Extraperitoneal Inguinal Hernia Repair? A Systematic Review. Surg. Endosc., 2007, 21 (2), 161–166. https://doi.org/10.1007/s00464-006-0167-4.

[14] Chen, L.-F. Applying Tissue and Mesh Combined Repair (TMC Repair) to Treat Adult Inguinal Hernia—A Study Based on 1,169 Cases. Front. Surg., 2022, 8. https://doi.org/10.3389/fsurg.2021.810212.

[15] Kohn, G. P.; Price, R. R.; Demeester, S. R.; Zehetner, J.; Muensterer, O. J.; Awad, Z.; Mittal, S. K.; Richardson, W. S.; Stefanidis, D.; Fanelli, R. D. Guidelines for the Management of Hiatal Hernia. Surgical Endoscopy. 2013, pp 4409–4428. https://doi.org/10.1007/s00464-013-3173-3.

[16] Jenkins, J. T.; O’Dwyer, P. J. Inguinal Hernias. BMJ, 2008, 336 (7638), 269–272. https://doi.org/10.1136/bmj.39450.428275.AD.

[17] Berle, M.; Dahlslett, K. H.; Kavaliauskiene, G.; Hoem, D. Internal Abdominal Hernia. Tidsskr. Nor. Laegeforen., 2017, 137 (16). https://doi.org/10.4045/tidsskr.17.0090.

[18] Białecki, J.; Pyda, P.; Antkowiak, R.; Domosławski, P. Unsuspected Femoral Hernias Diagnosed during Endoscopic Inguinal Hernia Repair. Adv. Clin. Exp. Med., 2021, 30 (2), 135–138. https://doi.org/10.17219/acem/130357.

[19] Berndsen, M. R.; Gudbjartsson, T.; Berndsen, F. H. [Inguinal Hernia - Review]. Laeknabladid, 2019, 105 (9), 385–391. https://doi.org/10.17992/lbl.2019.09.247.

[20] Kulacoglu, H. Current Options in Inguinal Hernia Repair in Adult Patients. Hippokratia, 2011, 15 (3), 223–231.

[21] Selçuk, D.; Kantarci, F.; Oğüt, G.; Korman, U. Radiological Evaluation of Internal Abdominal Hernias. Turk. J. Gastroenterol., 2005, 16 (2), 57–64.

[22] Mayagoitia González, J. C. [NO TITLE AVAILABLE]. Rev. Col. Bras. Cir., 2010, 37 (1), 004–005. https://doi.org/10.1590/S0100-69912010000100002.

[23] Aldoescu, S.; Patrascu, T.; Brezean, I. Predictors for Length of Hospital Stay after Inguinal Hernia Surgery. J. Med. Life, 2015, 8 (3), 350.

[24] Eklund, A.; Rudberg, C.; Leijonmarck, C.-E.; Rasmussen, I.; Spangen, L.; Wickbom, G.; Wingren, U.; Montgomery, A. Recurrent Inguinal Hernia: Randomized Multicenter Trial Comparing Laparoscopic and Lichtenstein Repair. Surg. Endosc., 2007, 21 (4), 634–640. https://doi.org/10.1007/s00464-006-9163-y.

[25] Yang, J.; Tong, D. N.; Yao, J.; Chen, W. Laparoscopic or Lichtenstein Repair for Recurrent Inguinal Hernia: A Meta-Analysis of Randomized Controlled Trials. ANZ J. Surg., 2013, 83 (5), 312–318. https://doi.org/10.1111/ans.12010.

[26] Iftikhar, N.; Kerawala, A. QUALITY OF LIFE AFTER INGUINAL HERNIA REPAIR. Pol. Przegl. Chir., 2021, 93 (3), 1–5. https://doi.org/10.5604/01.3001.0014.8218.

